# Identifying High-Need Patient Profiles That Respond to Intensive Care Management: Insights from the Camden Health Care Hotspotting RCT

**DOI:** 10.64898/2026.03.06.26347776

**Authors:** Sindhoora Prakash, Dawn Wiest, Hari Balasubramanian, Aaron Truchil

## Abstract

**Background:** Evaluations of complex care programs for high-need patients have yielded mixed results, and identifying patient subgroups may reveal differential intervention effects. This study aimed to use latent class analysis (LCA) to identify high-need patient subgroups within a randomized trial of the Camden Coalition’s Core Model and to examine differences in healthcare utilization and care team engagement.

**Methods & Findings:** We conducted a post-hoc exploratory analysis of a randomized controlled trial (ClinicalTrials.gov: NCT02090426) involving 780 adults aged 18 ― 80 years in Camden, New Jersey, who had multiple chronic conditions and frequent hospitalizations. Participants were assigned to receive multidisciplinary care management delivered by nurses, social workers, and community health workers for 3 ― 4 months following hospital discharge, or to usual care. LCA incorporated medical, behavioral, and social risk factors, as well as prior hospital utilization, to identify patient subgroups. Outcomes included inpatient readmissions and emergency department visits over two consecutive 6-month post-discharge periods, along with service hours delivered to intervention patients.

Four patient classes emerged: (1) Behavioral Health & Housing Instability, (2) Multi-system Medical Complexity, (3) Pulmonary Health & Substance Use, and (4) Lower Overall Complexity. In the second 6-month follow-up period, intervention patients had lower readmission rates compared with controls (-6.4 percentage points; 90% CI, −12.2 to −0.5). Subgroup differences included reduced readmissions in Class 4 and fewer emergency department visits in Class 1. Service intensity varied across classes, with Class 1 receiving the highest number of staff hours and Class 2 the lowest.

**Conclusion:** Patient segmentation revealed meaningful variation in healthcare utilization outcomes and care team engagement across high-need subgroups, suggesting that tailoring complex care interventions to specific patient profiles may improve program effectiveness and equity.

## INTRODUCTION

Healthcare systems face the critical challenge of managing a small subset of patients with frequent hospital admissions who account for disproportionate utilization and healthcare costs [1–3]. These individuals often present with intersecting medical, behavioral, and social needs that conventional care models struggle to address effectively [1, 4–5]. In response, complex care management programs have emerged as a promising strategy. By leveraging multidisciplinary teams to coordinate care, provide patient education, and support navigation of medical and social services, these programs aim to improve outcomes for this high-need, high-cost population [6].

Despite their promise, evaluations of complex care interventions have yielded mixed results, highlighting the challenge of achieving consistent effectiveness. Some studies report reductions in hospitalizations or emergency department use [7–11], while others find limited or no impact [12–15]. The landmark randomized trial of the Camden Coalition’s Core Model showed no overall effect on readmissions, raising questions about the generalizability of such interventions [15]. Post-hoc analyses, however, revealed substantial variation in treatment effects based on patient engagement, with social risk factors linked to lower engagement [16]. These findings suggest that the success of complex care programs may depend on identifying which patients are most likely to benefit and addressing the barriers to engagement and sustained participation.

Heterogeneity among frequent hospital users complicates both care delivery and evaluation. Regression to the mean and subgroup responses can obscure treatment effects [16–18], limiting the insight gained from traditional analyses. Emerging evidence shows that “one-size-fits-all” models often fail to meet the needs of socially complex patients [19,20]. For example, studies on engagement in behavioral health populations show that meaningful benefits often require extended timelines [21], while implementation research underscores the importance of flexible, multidisciplinary approaches to foster effective participation [22]. By identifying distinct patient profiles with shared medical, behavioral and social characteristics, segmentation approaches, such as latent class analysis (LCA), can reveal varying patterns in treatment effects that would otherwise be obscured in aggregate analyses [23–26].

In this post-hoc exploratory analysis of the Camden Core Model trial, we applied LCA to identify distinct patient subgroups and assessed hospital readmissions, emergency department visits and care team service utilization across groups. By incorporating clinical, behavioral, and social risk factors into subgroup identification, this study addresses limitations of claims-based risk stratification [20] and highlights how variation in patient needs shapes program effectiveness. Patients with complex health and social needs, including dual-eligible populations and those experiencing housing instability, often face disparities in access to coordinated care [27–29]. Identifying which subgroups are most likely to benefit, and which could face barriers to engagement, is therefore not only a matter of efficiency but also of equity. By uncovering subgroup-specific patterns, our analysis seeks to guide more equitable resource allocation and inform personalized interventions that advance precision medicine in complex care management.

## METHODS

### Design Overview, Setting, and Participants

The primary trial (ClinicalTrials.gov: NCT02090426, registered on March 17, 2014; see S2 Text and S3 Text for the original RCT analysis plan and extension) randomized 809 hospitalized patients between June 2, 2014, and September 13, 2017, to receive either care management services or usual care. The prespecified primary outcome was hospital readmissions within 180 days of discharge. Eligible participants (ages 18 ― 80 years) had at least two hospitalizations in a six-month period, and evidence of two or more chronic illnesses and social complexity in their medical records. Patients identified through a health information exchange were approached at Cooper University Hospital or Our Lady of Lourdes Hospital in Camden, New Jersey. After consenting and completing a pre-enrollment questionnaire, patients (n=809) were randomized via a secure, independent system into treatment (n=399) and control (n=401), after excluding 9 patients due to exceeding enrollment targets (Fig 1). The protocol was approved by Cooper University Healthcare, the National Bureau of Economic Research, and Our Lady of Lourdes Medical Center and is available with the original trial publication [15].

**Fig 1.**
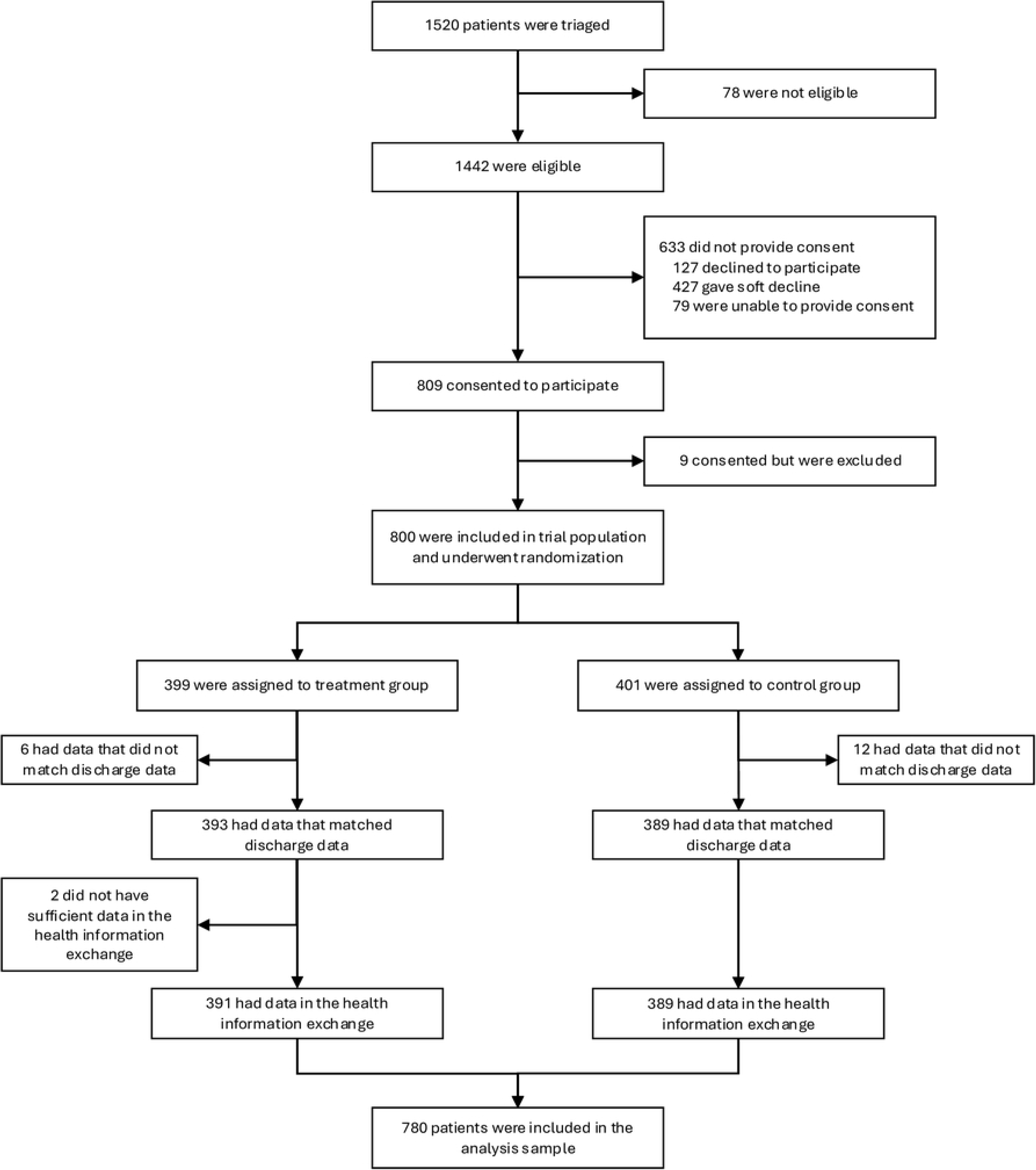
Randomized Control Trial Consort Diagram.

### Intervention

Care planning activities for patients in the treatment group began during the patient’s index hospitalization and continued after discharge through home and community visits by the Camden Coalition care team. A multidisciplinary team, comprising nurses (LPNs and RNs), social workers, community health workers and health coaches, collaborated with patients to assess needs and set goals. The team coordinated follow-up care; provided coaching in medication management, blood pressure monitoring, and other self-care activities; and assisted patients in applying for social services, behavioral health resources, and government benefits. Care team staff worked with patients until the patient was medically stable and graduated from the program, was lost to follow-up, died, or exited the intervention for another reason, such as relocation or declining further services.

### Outcomes

The prespecified primary outcome of hospital readmissions within 180 days of hospital discharge was expanded to include an additional 6-month follow-up period. In addition, emergency department visits in the two 6-month periods were analyzed. We also examined care team service utilization for intervention patients both up to and beyond the outcome date, defined by program graduation, or departure due to loss of contact or death. Utilization beyond the outcome date was included because patients in complex care programs may require continued support even after official program conclusion.

### Datasets

The primary program dataset from the randomized controlled trial (extracted April 2023) included 782 patients (393 intervention, 389 control), with key demographic, social and clinical characteristics, including a pre-computed comorbidity index. This dataset also captures granular engagement histories for intervention patients, with duration recorded for each intervention encounter. Additionally, we used hospital encounter data from the Camden Coalition Health Information Exchange (HIE) (extracted April 2023) rather than the original RCT claims data to obtain more detail on chronic conditions and emergency department visits. The HIE dataset covered the same 782 patients and included admission date, visit type (emergency department, inpatient, or outpatient), and ICD-9/10 diagnosis codes. Two intervention patients were excluded from our analysis: one had no inpatient visits in the HIE, and the other had no inpatient admission within six months prior to the index admission originally used to establish eligibility. This discrepancy likely reflects differences in how the HIE and claims systems classify and capture encounters, which can lead to occasional mismatches. Other possibilities include patients revoking consent or a missed linkage in the HIE. Because these data systems operate independently and with different assumptions and constraints, we can only speculate about the precise reason for the discrepancy.

### Defining Clinical, Social and Utilization Measures

Using ICD-9/10 codes directly in our analysis would make the analysis intractable due to the sheer number of unique codes present in the dataset. Hence, we make use of the Clinical Classification Software (CCS) from the Agency for Healthcare Research and Quality, which consolidates more than 14,000 ICD-9 diagnosis codes and 3,900 ICD-9 procedure codes into a smaller number of multi-level, clinically meaningful categories [30]. For this study, we used “Appendix C: Multi-Level Diagnoses” to map ICD-9 codes to CCS Levels 1 and 2. CCS Level 1 captures broad categories, typically by organ system, while Level 2 identifies major chronic conditions within each Level 1 category. ICD-10 codes in the dataset were converted to ICD-9 using the Centers for Medicare & Medicaid Services General Equivalence Mappings to enable CCS-based categorization [31].

We supplemented the CCS categories by creating several “custom” Level 2 groupings: HIV, Obesity, Nicotine Dependence, Chronic Kidney Disease, Cardiac Dysrhythmias, Coronary Artery Disease, and Stroke / Cerebral Ischemia, improving alignment with conditions measured in the RCT’s pre-enrollment survey. To better capture the role of smoking in respiratory health, we separated “Nicotine Dependence” from the broader “Alcohol-/Substance-Related Disorders” category. We also combined certain less prevalent conditions into broader categories, such as “Disorders of Mouth, Teeth, and Jaw (excluding dental),” “Other Gastrointestinal Disorders,” and “Other Infectious/Parasitic Diseases.” CCS Level 1 categories outside the scope of the trial were excluded. These included Neoplasms; Pregnancy, Childbirth, and the Puerperium; Skin and Subcutaneous Tissue Disorders; Congenital Anomalies; Conditions Originating in the Perinatal Period; Injury and Poisoning; Symptoms, Signs, and Ill-defined Conditions; Factors Influencing Health Status; Residual Codes; Unclassified Codes; and all E Codes. The final set of CCS-derived variables comprised 79 chronic medical and behavioral health conditions.

Additionally, we incorporated four social factors: housing status, marital status, education, and social support. These data were captured on a survey administered in person by Camden Coalition staff at hospital bedside prior to randomization. We also included measures of hospital utilization in the 6 months prior to study enrollment. The thresholds for the hospital utilization measures – 5 or more emergency department visits and 3 or more inpatient admissions – were determined based on clinical relevance and statistical distributions observed in the data. Post-enrollment hospital utilization outcomes were not used to compose the classes.

### Statistical Analysis

We conducted LCA to identify patient subgroups defined by pre-enrollment utilization patterns and clinical and social characteristics. LCA is a statistical method used to identify distinct, unobservable subgroups (latent classes) within a population based on patterns of association in observable characteristics or behaviors [32]. This technique maximizes within-class homogeneity and between-class heterogeneity, yielding a concise, interpretable set of classes. LCA has been successfully applied in healthcare research to uncover meaningful subgroups among patients with multiple comorbidities without imposing stringent constraints [33–42]. By employing LCA, researchers can gain insights into underlying patterns and relationships in complex datasets, potentially informing more targeted, cost-effective interventions and personalized care strategies.

We implemented LCA using the Stepmix v2.1.3 package in Python v3.7.10 [43]. The optimal number of classes was determined after assessing models with one to ten classes. Selection was guided by the Bayesian Information Criterion (BIC), which balances model fit and complexity, and the average posterior class membership probability, which reflects the model’s confidence in assigning individuals to classes [37]. Additionally, we considered the clinical interpretability of the classes; too few classes risked obscuring heterogeneity, while too many risked small, unstable groups. We ensured that each class included adequate representation from both intervention and control groups to support statistically valid comparisons of outcomes. Parameters for LCA were estimated using the expectation-maximization algorithm, which may converge to a local maximum depending on the randomized initial starting values [32]. Hence, we used an inbuilt feature in Stepmix to consider 100 unique initializations for each model to reduce the risk of suboptimal solutions.

To describe the resultant classes, we used demographic, clinical, and hospital variables, along with care team staff time for intervention patients only. Clinical complexity was quantified via the pre-computed Updated Charlson Comorbidity Index, which predicts mortality risk using weighted comorbidities [44]. Individual comorbidities are assigned weights from 0 (lowest risk) to 6 (highest risk). Binary features (e.g. gender, race) were analyzed using the Chi-squared contingency test, while discrete and continuous features (e.g. age, intervention hours) were analyzed using Welch’s test since the data does not follow a normal distribution and variances are unequal across latent classes [45]. Finally, we compared classes based on emergency department visits and inpatient admissions during two six-month periods after index hospital discharge. The differences in readmissions between the intervention and control groups were validated for statistical significance via two-sample proportion Z-test. Multiple testing adjustments were not applied because this study is explicitly designed to be exploratory and hypothesis-generating, and because there is no unambiguous basis for defining families of hypotheses across exposure groups, latent classes and time periods (See Section F in S1 Appendix for deeper analysis on adjustment for multiple testing) [46–48].

### Institutional Review Board (IRB) Approvals

The parent RCT was reviewed and approved by Cooper University Hospital Human Research Protection Program (Protocol #13-171, *Health Care Hotspotting: A Randomized Controlled Trial*; see S4 Text and S5 Text for approved protocol and amendment). The approval extends to this study, where the research activities are limited to data analysis. The analysis in this study was also reviewed and approved by the University of Massachusetts Human Research Protection Office (Protocol #3832, *Modeling the Impact of Care Interventions on Patients with Complex Medical and Social Needs*; see S6 Text – S9 Text for submitted protocol, amendment, and approval documentation).

## RESULTS

### Patient Characteristics

After excluding patients who did not have sufficient records in the HIE, the final analysis sample consisted of 780 patients: 391 in the intervention group; 389 in the control group. The mean age was 56.53 years; the majority identified as Black/African American (54.62%) or Hispanic/Latino (29.49%); one-half were women (50.51%) (Table 1). Additionally, 5.51% were employed and nearly 1 in 5 (18.59%) did not speak English. The patients had an average Updated Charlson Comorbidity Index of 2.42.

**Table 1.**
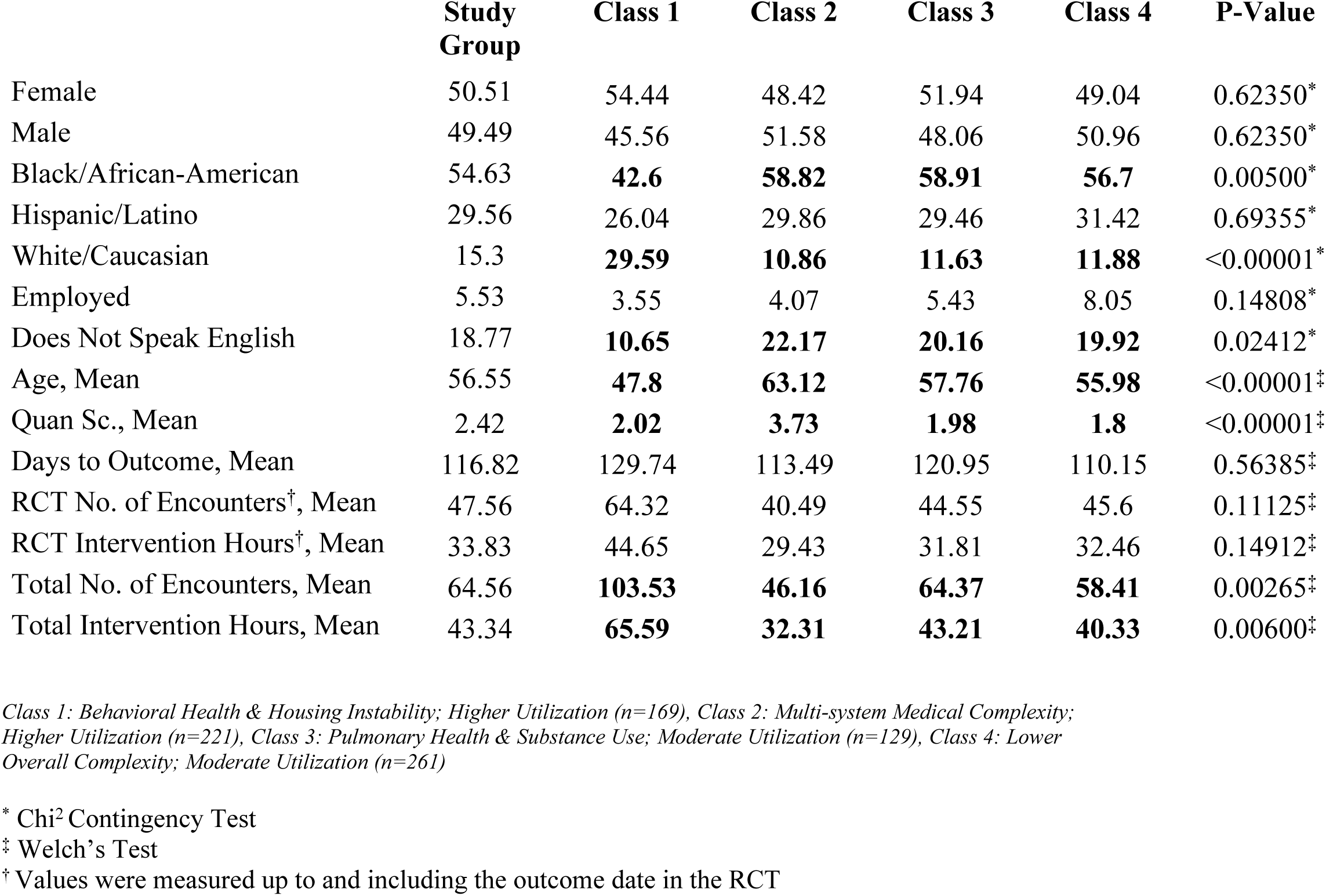
Demographics and intervention utilization characteristics for study population and latent classes.

### Segmentation by Past Hospital Utilization, Social Characteristics, and Clinical Conditions

We tested LCA models with one to ten classes and selected the four-class model as the best-fit solution based on the criteria described above (See Section A in S1 Appendix for statistical measures). Class sizes ranged from 16.54% to 33.46% of the study population, and the posterior probabilities of class membership ranged from 0.34 to 1.0, with mean values between 0.91 to 0.94 across classes. The model utilized 85 binary indicators (79 clinical conditions, 4 social determinants of health, and 2 hospital utilization measures). We identified distinctive features by examining the prevalence of each indicator within and across the classes. Fig 2 displays all model indicators for each class, shaded to reflect their prevalence. Indicators that define and label the groups are enclosed in color-coded boxes corresponding to each class. Hypertension and heart disease were common across all classes.

**Fig 2.**
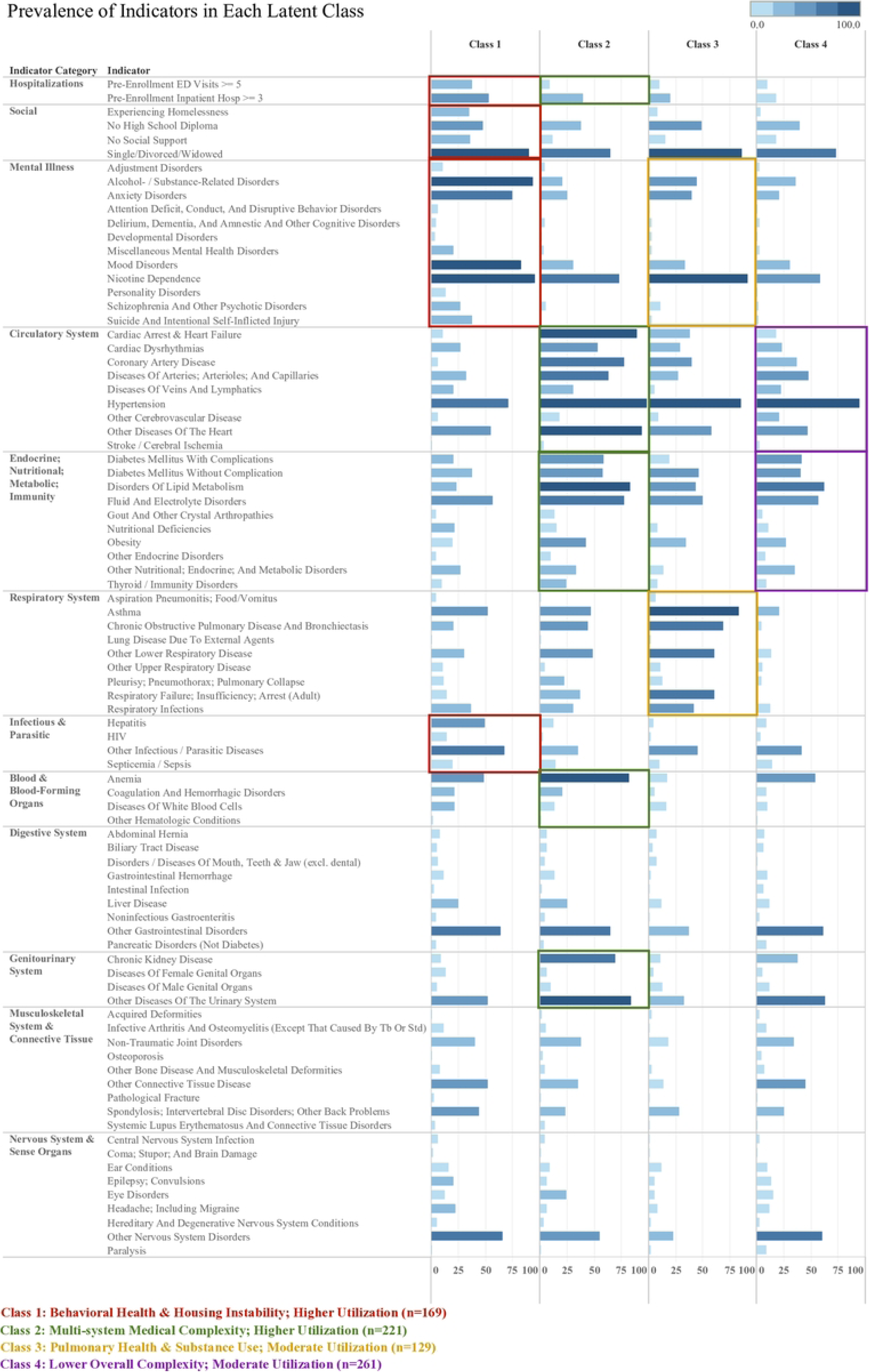
Pre-enrollment utilization, social, and clinical classifications within latent classes of complex patients enrolled in the RCT, with color coded boxes to highlight the indicators used for class labeling.

### Latent Class Characteristics

The analysis revealed four distinct patient classes with unique health and hospital utilization profiles:

- **Class 1: Behavioral Health & Housing Instability; Higher Utilization (n=169)** was characterized by the highest prevalence of mental health conditions, substance use disorders, HIV, hepatitis, and homelessness. This class had frequent emergency department visits and hospitalizations prior to enrollment.
- **Class 2: Multi-system Medical Complexity; Higher Utilization (n=221)** exhibited a broad spectrum of complex health conditions across multiple organ systems, particularly circulatory and endocrine/metabolic disorders. This class showed high rates of hospitalization and emergency department utilization.
- **Class 3: Pulmonary Health & Substance Use; Moderate Utilization (n=129)** was distinguished by respiratory conditions combined with co-occurring mental health issues, including anxiety and substance-use disorders. This class had a high prevalence of nicotine dependence comparable to Class 1.
- **Class 4: Lower Overall Complexity; Moderate Utilization (n=261)** demonstrated moderate circulatory system issues and some endocrine/metabolic disorders, but with lower overall disease complexity compared to the other three classes.

Classes 3 and 4 included fewer patients with 3 or more hospitalizations and 5 or more emergency department visits prior to enrollment compared to Classes 1 and 2.

### Demographics and Clinical Profiles by Class

Significant demographic differences existed across classes (Table 1). Mean age varied from 47.8 years in Class 1 to 63.12 years in Class 2, compared to the overall study population average of 56.55 years. Class 1 had a higher proportion of female patients (54.44%) and White/Caucasian patients (29.59%) compared to the overall study population (50.51% and 15.30%, respectively). Class 4 showed the highest employment rate (8.05% versus 5.53% overall). Medical complexity varied significantly across classes as measured by the comorbidity index. Class 2 had the highest average score (3.37), while Class 4 had the lowest (1.8), compared to the overall study average of 2.42.

### Intervention Utilization Patterns by Class

Among intervention group patients, utilization of care team services beyond the outcome date varied significantly across classes (P<0.001). Class 1 had the highest average service hours until outcome (44.65 hours) and beyond outcome (65.59 hours), exceeding overall averages of 33.83 and 43.34 hours, respectively. Despite higher medical complexity, Class 2 showed the lowest average utilization both until outcome (29.43 hours) and beyond outcome (32.31 hours).

### Post-enrollment Healthcare Utilization Outcomes

Analysis of the full study population showed no statistically significant differences in hospital readmission or emergency department visit rates between intervention and control groups during the first six-month post-enrollment period (Table 2). In the second six-month period, intervention patients demonstrated a trend towards reduced inpatient readmission rates compared to control patients (-6.37, 90% CI: −12.15 to −0.54, P=0.073). Class-specific analyses revealed additional patterns in the second six-month period. Class 4 intervention patients had significantly lower inpatient readmission rates compared to controls (−11.18, 90% CI: −20.86 to −1.2, P=0.067). Class 1 intervention patients showed reduced emergency department visit rates compared to controls (−13.51, 90% CI: −25.28 to −1.35, P=0.07). In the pre-enrollment period, Class 4 intervention patients had higher emergency department visit rates compared to their control counterparts (11.16%, 90% CI: 1.08 to 20.93, P=0.07), but this difference was not observed in either post-enrollment period. No other classes demonstrated statistically significant differences in readmission or emergency department visit rates across the study periods.

**Table 2:**
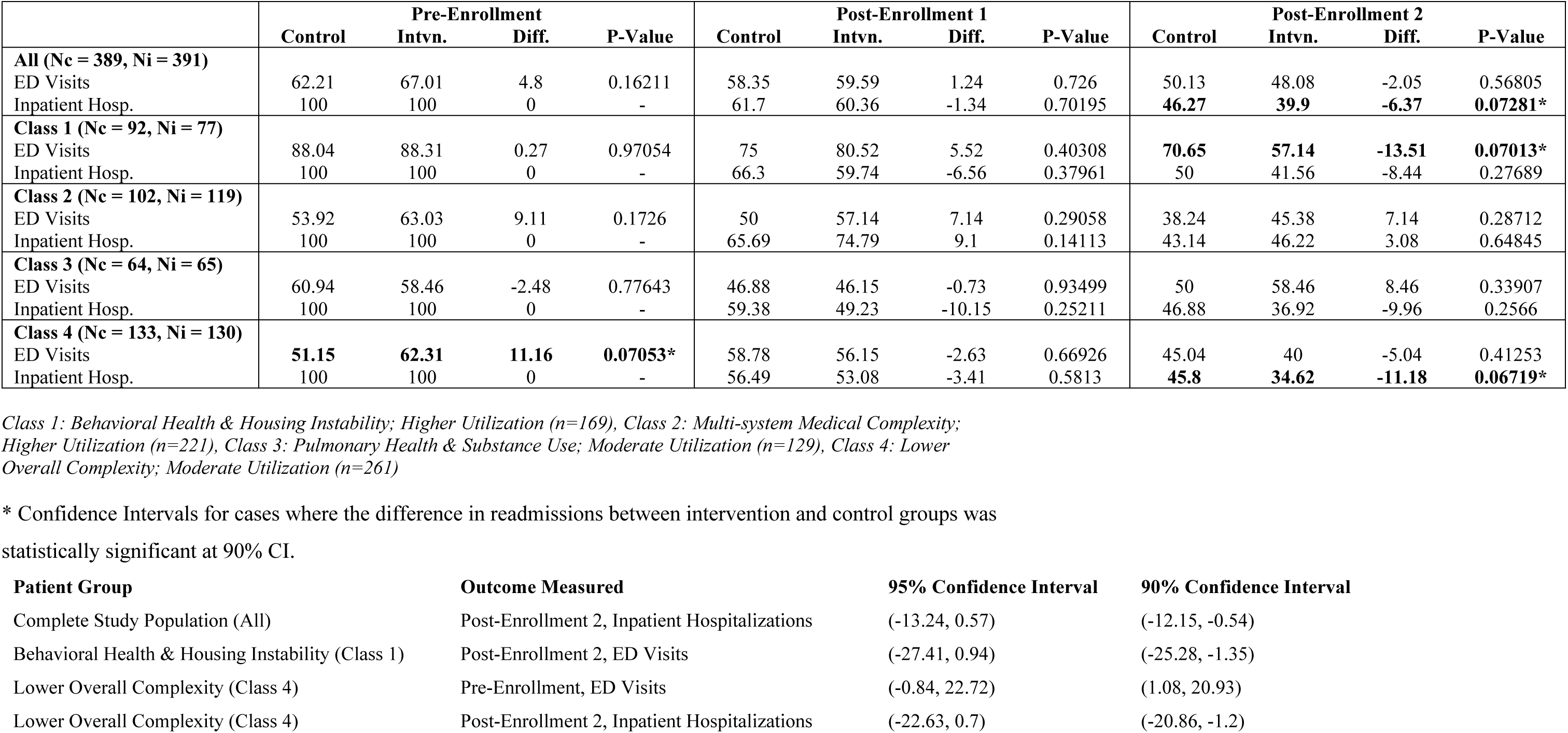
Emergency department and inpatient admission rates for intervention and control group patients within the study population and each class for the 6-month pre-enrollment and two post-enrollment periods (P-values for two-sample proportion Z-test).

## DISCUSSION

This study identified four distinct classes of high-need patients using LCA, each with markedly distinct clinical and social profiles and utilization patterns. By integrating clinical, behavioral, and social risk factors, this analysis extends prior evaluations of the Camden Core Model trial by helping explain why uniform care management approaches have yielded mixed results across complex populations. Our findings reinforce the critical insight raised in the introduction: heterogeneity among high utilizers cannot be addressed effectively by “one-size-fits-all” care models.

### Heterogeneity and Regression to the Mean

Consistent with prior concerns about regression to the mean and obscured subgroup effects [16–18], our analysis demonstrates that average treatment effects mask important variations. While the parent trial found no overall impact on readmissions, we observed limited and subgroup-specific differences in utilization. Among Behavioral Health & Housing Instability patients, intervention participants showed initially higher ED use but a subsequent decline relative to controls by the second follow-up period (57% vs. 71%, P=0.07), suggesting delayed improvement. Multi-system Medical Complexity patients remained persistently high utilizers with no significant differences by study arm. Pulmonary & Substance Use patients showed stable patterns of ED and inpatient use across groups, with no evidence of intervention benefit. Finally, Lower Complexity patients demonstrated reductions in inpatient hospitalizations at the second follow-up (35% vs. 46%, P=0.067), despite only modest baseline risk. These findings suggest that improvements emerged in selected groups, but not consistently across the population, underscoring the importance of tailoring interventions to subgroup needs.

### Addressing Socially Situated Risk

Our results underscore the limitations of traditional risk stratification based on claims or medical data alone. As Nong and Adler-Milstein note, algorithmic scores often overlook socially situated risks such as isolation, behavioral health needs, or housing instability [20]. By incorporating social and behavioral factors alongside clinical data, our LCA approach identified subgroups that are both empirically robust and clinically interpretable. Importantly, the signal of improvement among Lower Complexity patients—who might not otherwise be flagged by risk scores—suggests that stratification approaches that combine medical and social dimensions can better capture who stands to benefit from support.

### Engagement, Implementation and Program Design Implications

Differences in service utilization suggest that engagement variation may help explain differential effects, echoing post-hoc analyses of the Camden trial showing that treatment effects varied by patient engagement [49]. Behavioral Health & Housing Instability patients may have faced early barriers to engagement, with improvements emerging later, indicating a need for sustained outreach and high-intensity support. Multi-system Medical Complexity patients showed persistently high hospital utilization despite intervention, suggesting a need for more specialized care models. Pulmonary & Substance Use patients showed no measurable benefit, implying their patterns are less responsive to short-term interventions and that targeted supports for substance use or pulmonary health may be more effective. Finally, Lower Complexity patients benefited through reduced inpatient admissions, showing that lower-risk individuals can benefit from lighter-touch supports that prevent escalation. Tailoring program design and intensity to these patterns may respond directly to concerns about equity and efficiency raised in the introduction.

### Equity and Access

The demographic differences across subgroups raise equity considerations. Patients in the Behavioral Health & Housing Instability class were substantially younger (mean age 48 vs. 57 overall, P<0.001) and disproportionately White (30% vs. 15% overall, P<0.001), highlighting that vulnerability is shaped less by age or medical burden and more by behavioral health and housing instability. By contrast, patients in the Multi-system Medical Complexity and Pulmonary & Substance Use classes were older (mean ages 63 and 58, respectively), included more patients identifying as Black (∼59% in both vs. 55% overall, P=0.005), and more likely to be non-English speaking (22% and 20%, respectively vs. 19% overall, P=0.024), suggesting that these patients face both clinical and sociocultural barriers. Despite these challenges, analyses show improvements emerged unevenly. Excluding patients due to perceived barriers—language, age, or social complexity—would risk exacerbating disparities in access to coordinated care. At the same time, the favorable outcomes observed in the Lower Complexity group underscore that equity-focused strategies must avoid overlooking lower-risk patients, who may fall through the cracks of programs focused only on the most visibly high-need. These findings suggest that equity and effectiveness are inseparable: program resource allocation across subgroups directly shapes both outcomes and disparities.

### Limitations and Future Research

Several important limitations warrant consideration. First, our outcome measure of readmission rates presents inherent challenges in distinguishing planned from unplanned readmissions and preventable from non-preventable cases, which complicates assessments of the intervention’s direct effect [50–52]. Second, the reverse directionality in inpatient readmission outcomes in the first post-intervention period between the original RCT and our study may be explained by differences in the analytic sample and data used, that is, the exclusion of two intervention patients and the use of claims vs. HIE (See Section E in S1 Appendix for a comparison). Hence, these findings should be viewed as exploratory, aimed at generating insights into how intervention effects may vary across different patient profiles rather than a definitive reflection of the original RCT’s results. Third, our use of custom chronic condition groupings at the CCS Level 2 level, while designed to enhance analytical precision for specific conditions and address coding inconsistencies, may limit comparability with other studies using standard groupings. Finally, the LCA algorithm can generate multiple statistically valid solutions with varying numbers of classes. Although the Camden Coalition’s Chief Medical Officer initially favored a simpler two-class model, we concluded that the four-class solution better captured population heterogeneity (See Section C in S1 Appendix for 2- & 3-class models). This underscores that while clinical input is valuable, it may not always align with statistical criteria or the analytic goal of identifying nuanced subgroups. The selection process remains inherently subjective despite statistical guidance.

Future work should explore: 1) optimal outreach and engagement strategies for socially complex patients, 2) sustainability of intensive interventions for persistently high utilizers, 3) step-down protocols for more stable patients who may not require ongoing high-intensity services, and 4) transitional or preventive approaches for lower complexity patients, who nonetheless demonstrated measurable benefit. Cost-effectiveness analyses are also needed to determine how segmentation-informed approaches can improve both efficiency and equity. Importantly, healthcare systems should replicate segmentation analyses in their own patient panels to ensure context-specific applicability.

## Conclusion

By uncovering distinct subgroups and trajectories within a heterogeneous trial population, this study demonstrates how segmentation can illuminate variation obscured in aggregate analyses and address concerns about regression to the mean. Our findings suggest that effectiveness depends less on producing large average effects across all patients and more on identifying where intervention signals emerge and matching resources accordingly. Integrating LCA with implementation science frameworks, and eventually AI-enabled precision medicine, offers a path forward for healthcare systems: moving beyond one-size-fits-all models toward equitable, evidence-based, and patient-centered strategies that improve outcomes for populations with the most complex needs.

## Data Availability

The datasets used in this study contain detailed demographic and clinical information for patients within a specific geographical region. Due to the potentially identifiable nature of these datasets, and the applicable privacy and ethical restrictions, they are not publicly available. However, data with the minimum variables required to replicate the results in this study may be made available upon request. Please direct data-related queries to the corresponding author(s).

## ACKNOWLEDGEMENTS

We thank the Camden Coalition care team for their dedication to the patients they serve. We are grateful to Jubril Oyeyemi, MD (Camden Coalition), for feedback on clinical validity of subgroups, and to Kathleen Noonan, JD (Camden Coalition) and the Camden Coalition care team for their critical feedback on the manuscript’s contents. No compensation was provided.

## SUPPORTING INFORMATION

S1 Appendix. Supplemental Material.

S2 Text. Original RCT NCT02090426 Analysis Plan.

S3 Text. Original RCT NCT02090426 Analysis Plan Extension.

S4 Text. Cooper University Hospital Human Research Protection Program IRB # 13-171 Study Protocol.

S5 Text. Cooper University Hospital Human Research Protection Program IRB # 13-171 Amendment.

S6 Text. University of Massachusetts Human Research Protection Office IRB # 3832 Study Protocol.

S7 Text. University of Massachusetts Human Research Protection Office IRB # 3832 Study Protocol Approval Letter.

S8 Text. University of Massachusetts Human Research Protection Office IRB # 3832 Amendment.

S9 Text. University of Massachusetts Human Research Protection Office IRB # 3832 Amendment Approval Letter.

S10 Checklist. CONSORT 2025 checklist of information to include when reporting a randomised trial.

